# Surveillance for Opioid-Associated Amnestic Syndrome in Canada, 2010-2022

**DOI:** 10.1101/2024.10.24.24315474

**Authors:** Jed Barash

**Affiliations:** Department of Medicine, Veterans Home, Chelsea, Massachusetts

## Abstract

**Objectives:** A single diagnostic code could be used to perform surveillance for opioid-associated amnestic syndrome (OAS) in healthcare datasets on a national scale.

**Methods:** A request was submitted to search the Discharge Abstract Database (DAD) and the National Ambulatory Care Reporting System (NACRS) in Canada for the ICD-10-CA code, F11.6 (mental and behavioural disorders due to use of opioids, amnesic syndrome) during fiscal years (FY) 2010-2011 through 2022-2023. The annual total of encounters using this code was determined and crude rates per million were calculated for the Canadian population represented.

**Results:** National counts from DAD and NACRS combined ranged from <5 to 14 annually. Rates per million for available years fell between 0.17 and 0.48. For available data through FY 2015-2016, the mean number of annual combined encounters nationally was 6.3; for those years after, the mean was 10. The mean rate per million was 0.23 and 0.35 for these two periods, respectively.

**Discussion:** This study represents the first effort to conduct surveillance for OAS on a national scale and suggests that the condition is relatively rare. Future efforts to validate the coded diagnosis of OAS with confirmed cases will help determine its value as a surveillance tool.

## Introduction

Recent years have witnessed the emergence of an acute anterograde amnestic syndrome linked to injury of the hippocampi as evident on brain imaging and a history of opioid misuse.^1,2^ Cases of this opioid-associated amnestic syndrome (OAS) have been identified internationally, including in the United States^3-5^ and Canada.^6-8^ OAS may be diagnosed after presentation to the emergency department for evaluation of confusion or after extubation in the setting of an admission for overdose; memory loss may persist for months or longer.^1^

Published reports suggest that OAS is rare, but efforts to estimate its frequency have been limited. In the United States, a prior study of OAS in Massachusetts examined traditional reporting through health care providers as well as syndromic surveillance restricted to both free-text data and a series of related codes from emergency department (ED) visits.^9^

The potential for surveillance in Canada is strengthened by: 1) a single, existing code (F11.6, mental and behavioural disorders due to use of opioids, amnesic syndrome) in the *International Statistical Classification of Diseases and Related Health Problems, Tenth Revision, Canada* (ICD-10-CA) to identify possible cases of OAS and 2) long-standing ED and inpatient data captured at a national level. This study aimed to leverage these features to perform surveillance for possible cases of OAS in Canada.

## Methods

A request was submitted to the Canadian Institute for Health Information (CIHI) to search the Discharge Abstract Database (DAD) and the National Ambulatory Care Reporting System (NACRS) for the ICD-10-CA code, F11.6 (mental and behavioural disorders due to use of opioids, amnesic syndrome). Though this code has been in existence since the implementation of the ICD-10-CA in 2001, the fiscal years (FY) 2010-2011 through 2022-2023 were selected as the interval of study based on: 1) the establishment of a baseline prior to the earliest known OAS cases reported in the literature (2012)^1^ and 2) the most recent year of accessible data.

DAD includes encounters at the acute inpatient level of care, while NACRS includes those for the emergency department. For both DAD and NACRS, the search involved data from all submitting provinces, with the understanding that Quebec has not participated in this process during the period of investigation. Submission was not mandated for all facilities. Based on CIHI’s privacy and confidentiality protocols, non-zero counts less than 5 were suppressed.

Given these protocols and the relative rarity of the diagnostic code of interest, annual counts nationally from DAD and NACRS were combined into a single total.

Crude rates of encounters assigned the F11.6 code per million were calculated for each year through FY 2022-2023 using Canadian population estimates, excluding Quebec.^10^ Fiscal years start on April 1^st^ and end on March 31^st^ the following year.

### Standard Protocol Approvals, Registrations, and Patient Consents

This study used deidentified data and did not require institutional review board approval. DAD and NACRS are national datasets that do not obtain consent from participants.

### Data Availability

Qualified researchers may request access to all deidentified DAD and NACRS registry data used for this study through CIHI.

## Results

Available annual combined counts nationally from DAD and NACRS ranged from <5 in FY 2010-2011 and FY 2013-2014 to 14 in FY 2019-2020 (Table 1). Rates per million for years without suppressed counts fell between 0.17 (FY 2020-2021) to 0.48 (FY 2019-2020).

**Table.** Annual counts and rates of *ICD-10-CA* code F11.6 (mental and behavioural disorders due to use of opioids, amnesic syndrome) in Canada* from Discharge Abstract Database and National Ambulatory Care Reporting System combined, fiscal years 2010-2011 through 2022-2023.

| Fiscal Year | Total Count | Rate (per million) |
| --- | --- | --- |
| 2010-2011 | N/R | N/R |
| 2011-2012 | 5 | 0.19 |
| 2012-2013 | 9 | 0.34 |
| 2013-2014 | N/R | N/R |
| 2014-2015 | 5 | 0.18 |
| 2015-2016 | 6 | 0.22 |
| 2016-2017 | 11 | 0.40 |
| 2017-2018 | 10 | 0.36 |
| 2018-2019 | 11 | 0.39 |
| 2019-2020 | 14 | 0.48 |
| 2020-2021 | 5 | 0.17 |
| 2021-2022 | 11 | 0.37 |
| 2022-2023 | 8 | 0.27 |

Abbreviations: ICD-10-CA: International Statistical Classification of Diseases and Related Health Problems, Tenth Revision, Canada; N/R: Not reported (non-zero counts less than 5 are suppressed by protocol, thus rates cannot be reported either)
\*Excluding province of Quebec, which does not report

For years in which data was not suppressed between FY 2010-2011 and 2015-2016, the mean number of annual combined encounters nationally was 6.3; for those years after, the mean was 10. The mean rate per million was 0.23 and 0.35 for these two periods, respectively.

## Discussion

This study represents the first published effort to conduct surveillance of OAS on a national scale. Using administrative datasets and a single ICD-10 code, Canadian data suggests that the condition is relatively rare, consistent with prior evidence.^9^ With the exception of FY 2020-2021 (potentially attributable to the COVID-19 pandemic), there has generally been an increase in encounter counts and the associated crude rate since FY 2016-2017, following an acceleration in the Canadian epidemic of illicit synthetic opioid use^11^ and preceding the earliest published reports of Canadian OAS cases.^6,7^ The calculated differences in mean numbers between these periods are, in fact, conservative, given data suppression during both FY 2010-2011 and 2013-2014.

Systemic factors with data collection, primarily involving non-mandatory participation by facilities, may have led to under detection of coded encounters. By contrast, the possibility that encounters may not represent unique patients or cases of OAS altogether would lead to overestimates of frequency. Future efforts to validate the coded diagnosis of OAS with confirmed cases will help determine its value as a surveillance tool, particularly in the United States, where OAS can now be reported based on inclusion terms added to the ICD-10 in 2023 under codes for opioid dependence (F11.288) and opioid use (F11.988).

## Acknowledgments

Parts of this material are based on data and information provided by the Canadian Institute for Health Information. However, the analyses, conclusions, opinions, and statements expressed herein are those of the author and not necessarily those of the Canadian Institute for Health Information.

The author expresses appreciation to Dr. Alfred DeMaria for his critical review of the manuscript.

